# Categorical and Dimensional Alterations Along Two Principal Cortical Gradient Axes Across the Schizophrenia-Bipolar Spectrum

**DOI:** 10.64898/2026.06.30.26356921

**Authors:** Asia Ferrari, Bin Wan, Judith Kabbeck, Amin Saberi, Stefan Kaiser, Valeria Kebets, Clara Moreau, Paul M. Thompson, Theo G.M. Van Erp, Jessica A. Turner, B.T. Thomas Yeo, Boris C. Bernhardt, Sofie L. Valk, Matthias Kirschner

## Abstract

**Background and Hypothesis:** Schizophrenia (SZ) and bipolar disorder (BD) share overlapping yet distinct clinical profiles and system-wide brain alterations. Macroscale functional connectivity gradients capture principal axes of cortical organization, including the separation of unimodal and transmodal systems, offering a low-dimensional lens on individual differences in brain architecture. Whether these axes reflect shared or diagnosis-specific variation across the SZ-BD spectrum is unknown.

**Study Design:** Using resting-state fMRI from 187 adults (110 HC, 37 SZ, 40 BD) from the UCLA Consortium for Neuropsychiatric Phenomics, we derived individual low-dimensional gradients and applied three analyses: case-control comparisons at both the cortical network and subcortical region-of-interest level, Partial Least Squares (PLS) regression linking gradients to clinical phenotypes, and individual-level similarity indices (SI-PLS) positioning participants within a gradient–behaviour space.

**Study Results:** While the gradient structure (G1: visual-somatomotor and G2: unimodal-transmodal) was preserved across groups, patient groups showed greater deviations along both axes. Network analyses revealed transdiagnostic frontoparietal compression in G2, alongside disorder-specific effects: visual pole contraction and subcortical amygdala displacement in SZ, and somatomotor displacement in BD. PLS identified a BD-associated profile of preserved gradient architecture and lower symptom burden, contrasting with an SZ-associated profile of greater cognitive impairment and symptom severity. SI-PLS scores placed SZ and BD in distinct regions of a shared two-dimensional neural space, with HC between them.

**Conclusions:** Differences across the SZ–BD spectrum organize along two principal axes, revealing transdiagnostic alterations in higher-order association systems alongside disorder-specific sensory signatures. These findings support a multi-axis dimensional framework for understanding clinical heterogeneity in psychosis.

## Introduction

Schizophrenia (SZ) and bipolar disorder (BD) are among the most severe psychiatric conditions, characterized by their chronic disease course, heterogeneous symptoms expression, and substantial impact on functioning and quality of life^1–3^. Although traditionally conceptualized as distinct diagnoses^4^, both disorders share substantial overlap in symptom expression, cognitive impairment, genetic risk, and neurobiological substrates, including convergent alterations in dopaminergic and motivational circuitry^5–13^. Positive and negative symptoms are central to SZ^14^, but also recognized in BD^15,16^. Affective disturbances are hallmarks of BD, but frequently co-occur in SZ, and cognitive impairment, traditionally described as a distinguishing feature of SZ, is similarly observed, with variable severity, in BD^17–19^. The shared clinical manifestations of SZ and BD, alongside the heterogeneity within each diagnostic category, highlight limitations of categorical diagnostic boundaries, and motivate dimensional and transdiagnostic frameworks^20,21^.

Both disorders are further characterized by widespread structural and functional brain alterations, with shared as well as disorder-specific neural correlates. Large-scale meta-analyses document robust brain-wide morphometric disruptions with considerable cross-disorder overlap^6,22–24^. Multivariate analyses further demonstrate systematic associations between distributed structural and functional neuroimaging features and dimensional clinical phenotypes across SZ-BD^25–27^. At the large-scale functional network level, both disorders implicate higher-order systems, including the default mode, frontoparietal, and salience networks, though with partly divergent patterns, with BD preferentially involving anterior default mode and frontoparietal networks and SZ the posterior default mode and salience networks^28–33^.

Functional networks can be understood as part of a broader cortical organization, in which connectivity varies smoothly along continuous spatial axes, functional gradients, that arrange cortical regions into brain-wide hierarchies. The principal gradient runs from sensorimotor regions to transmodal association areas, while a secondary gradient separates visual from somatomotor regions^34–36^. These gradients can be estimated for each individual separately, are low-dimensional and biologically grounded, and recapitulate fundamental features of cortical organization including microstructure, neurodevelopment, and intrinsic dynamics^37–39^. Whether this hierarchical organization is itself altered across the SZ–BD spectrum, beyond focal network alterations, remains an open question^40–42^.

Prior work applying this framework to SZ reported compression along the principal sensorimotor-to-transmodal axis, across both early and chronic stages^43–45^. In contrast, Holmes et al. (2023)^46^ found the principal gradient largely preserved while observing disruptions along the secondary visual-to-sensorimotor gradient only in established SZ, in a sample that partially overlaps with our cohort. To our knowledge, no prior study has directly examined gradient organization across the SZ–BD spectrum. Specifically, it remains unknown to what extent gradient case-control effects are disease-specific or cross-disorder, and how individual-level gradient organization maps onto dimensional clinical variation across both diagnoses.

Here we address these questions by leveraging clinical and resting-state fMRI data from a transdiagnostic SZ–BD cohort^47^ and pursuing three complementary aims: (i) identifying categorical case-control effects on the two principal cortical gradients to distinguish disease-specific from cross-disorder alterations; (ii) examining whether gradient organization maps onto dimensional clinical-cognitive phenotypes across the SZ–BD spectrum, using multivariate PLS; and (iii) quantifying the individual-level expression of these gradient–behaviour dimensions, enabling continuous positioning of individual participants within the SZ-BD spectrum neural space. We hypothesize that (i) SZ and BD would show overlapping yet partially dissociable gradient alterations, (ii) that clinical variance would distribute across both gradient axes with diagnosis-specific contributions rather than collapsing onto a single dimension, and (iii) that individual position within this dimensional gradient-behaviour space would carry meaningful information beyond diagnostic category.

## Materials and Methods

### Participants and Study Design

We analyzed resting-state functional magnetic resonance imaging (rs-fMRI) data from a transdiagnostic cohort of 187 adults originally recruited for the UCLA Consortium for Neuropsychiatric Phenomics^47^, a broader transdiagnostic study that additionally included individuals with attention-deficit/hyperactivity disorder and schizoaffective disorder. The full cohort was previously analyzed across all diagnostic groups by Kebets et al. (2019)^48^ using whole-brain resting-state functional connectivity; here we restrict analyses to the schizophrenia–bipolar spectrum and adopt a cortical-gradient framework. The final sample comprised 110 healthy controls (HC), 37 individuals with schizophrenia (SZ), and 40 individuals with bipolar disorder (BD), with diagnoses confirmed using the Structured Clinical Interview for DSM-IV. Participants were selected based on the availability of preprocessed rs-fMRI data alongside comprehensive clinical and cognitive phenotyping, as described in Kebets et al. (2019)^48^. The UCLA Institutional Review Board approved all study procedures, and all participants provided written informed consent.

### Clinical and Cognitive Phenotyping

Clinical symptomatology was assessed using the Scale for the Assessment of Positive Symptoms (SAPS)^49^, the Scale for the Assessment of Negative Symptoms (SANS)^50^, the Young Mania Rating Scale (YMRS)^51^, and the 21-item Hamilton Depression Rating Scale (HAMD-21)^52^. Two domain scores were derived from individual SAPS item scores: positive symptoms, comprising delusions and hallucinations, and disorganization, comprising bizarre behaviour and positive formal thought disorder. For negative symptoms, two composite domain scores were computed: diminished expression, averaging blunted affect and alogia, and amotivation, averaging avolition/apathy and anhedonia/asociality^53^. Manic and depressive symptom severity were indexed by total YMRS and HAMD-21 scores, respectively. Cognitive performance was assessed across four domains using the standardized neuropsychological battery administered within the UCLA Consortium for Neuropsychiatric Phenomics^48,54^. Domain composite scores were computed by z-scoring and averaging test scores within each domain: attention and working memory (Digit Span Forward, Backward, and Sequencing; Letter–Number Sequencing), verbal learning (short- and long-delay free recall), reasoning (Matrix Reasoning), and vocabulary (Vocabulary). All composite scores were sign-flipped such that higher values reflect greater cognitive impairment.

### Statistical Comparisons of Sample Characteristics

Group differences in continuous demographic and cognitive variables were assessed using one-way analysis of variance (ANOVA), with Benjamini-Hochberg false discovery rate (FDR)^55^ corrected pairwise Welch’s t-tests for *post hoc* comparisons. Sex differences across groups were evaluated using a chi-squared test. Clinical variables were compared between patient groups only using Welch’s two-sample t-tests. Demographic and clinical characteristics are summarized in **Table 1**.

**Table 1.** Sample Characteristics. Values are mean ± SD for continuous variables; n (%) for sex. Cognitive composite scores are sign-flipped such that higher values indicate greater impairment. Abbreviations: HAMD-21, Hamilton Depression Rating Scale; N.A., not applicable; WM, working memory; YMRS, Young Mania Rating Scale; HC, Healthy Controls; SZ, Schizophrenia; BD, Bipolar Disorder. *p < .05 **p < .01 ***p < .001.

| Variable | HC<br>(N=110) | SZ (N=37) | BD (N=40) | Statistic | p-value<br>FDR<br>corrected | HC vs SZ<br>(p-value) | HC vs BD<br>(p-value) | SZ vs BD (p-value) |
| --- | --- | --- | --- | --- | --- | --- | --- | --- |
| **DEMOGRAPHICS** |  |  |  |  |  |  |  |  |
| **Age (years)** | 31.3 ± 8.7 | 35.5 ± 9.1 | 34.8 ± 9.2 | F=4.4 | 0.01* | 0.04* | 0.1 | 0.7 |
| **Sex (% male)** | 57 (52%) | 29 (78%) | 23 (58%) | $\chi^2=8.1$ | 0.02* | 0.01** | 0.6 | 0.08 |
| **CLINICAL DATA** |  |  |  |  |  |  |  |  |
| **HAMD-21** | N.A. | 11.9 ± 9.4 | 13.5 ± 10.0 | t=-0.7 | 0.5 | N.A. | N.A. | 0.5 |
| **YMRS** | N.A. | 8.9 ± 7.7 | 10.8 ± 10.5 | t=-0.9 | 0.4 | N.A. | N.A. | 0.4 |
| **Positive**<br>**Symptoms** | N.A. | 4.6 ± 2.7 | 0.8 ± 1.4 | t=7.5 | <.001*** | N.A. | N.A. | <.001*** |
| **Disorganization** | N.A. | 2.4 ± 2.3 | 1.4 ± 1.7 | t=2.1 | 0.04* | N.A. | N.A. | 0.04* |
| **Amotivation** | N.A. | 5.0 ± 2.8 | 3.6 ± 2.4 | t=2.4 | 0.02* | N.A. | N.A. | 0.02* |
| **Diminished**<br>**Expression** | N.A. | 2.2 ± 2.3 | 0.8 ± 1.3 | t=3.4 | 0.002** | N.A. | N.A. | 0.002** |
| **COGNITIVE Impairments (z-score)** |  |  |  |  |  |  |  |  |
| **Attention / WM** | -0.2 ± 0.7 | 0.6 ± 0.7 | 0.1 ± 0.7 | F=19.7 | <.001*** | <.001*** | 0.03* | 0.002** |
| **Verbal Learning** | -0.4 ± 0.7 | 0.9 ± 0.7 | 0.3 ± 0.8 | F=47.7 | <.001*** | <.001*** | <.001*** | 0.001** |
| **Reasoning** | -0.3 ± 0.9 | 0.8 ± 1.0 | -0.0 ± 0.9 | F=20.4 | <.001*** | <.001*** | 0.2 | <.001*** |
| **Vocabulary** | -0.3 ± 0.8 | 0.9 ± 0.9 | -0.2 ± 1.0 | F=25.7 | <.001*** | <.001*** | 0.7 | <.001*** |

### Functional Connectome and Macroscale Gradient Estimation

We utilized preprocessed resting-state data derived from Kebets et al., 2019^48^. Detailed preprocessing steps and parameters are reported in the **Supplementary Materials**. Subject-specific functional connectomes were constructed by calculating Pearson correlation coefficients between the averaged blood-oxygen-level-dependent (BOLD) time series of 400 cortical parcels defined by the Schaefer atlas^56^. Then Fisher *r*-to-*z* was employed on the connectome matrix. Macroscale cortical gradients were estimated for each participant using diffusion map embedding implemented via the BrainSpace toolbox^35^. Following thresholding of individual connectivity matrices to 90% row-wise sparsity, we applied a normalized angle kernel and performed diffusion embedding (α=0.5) to capture the dominant axes of connectome variance while preserving global geometric properties. Individual gradients were aligned to a reference template derived from HC-group-level using Procrustes rotation to maximize intersubject topographic correspondence. HC-group-level gradients were calculated based on the mean connectivity matrix across controls. The first two principal gradients (G1-G2) were retained for all downstream statistical models^57,58^. For each participant, Procrustes disparity was computed as a unit-less index of individual deviation from the normative gradient configuration after optimal rigid alignment (**see Supplementary Methods**).

### Parcel-wise and Network-Level Gradient Modelling

To elucidate case-control differences on macroscale functional architecture, we conducted comprehensive parcel-wise and network-level linear modelling. For parcel-wise mapping, we fit independent linear regression models for each of the 400 regions in each of the gradients G1 and G2. Gradient score served as the dependent variable, with diagnostic contrasts (HC vs. SZ, HC vs. BD, and HC vs. patients) as primary predictors, adjusting for age and sex. We additionally evaluated functional disorganization at the systems level by averaging parcel gradient scores within seven canonical resting-state networks (Visual, Somatomotor, Dorsal Attention, Salience, Limbic, Frontoparietal, and Default Mode)^56^. For each gradient, covariate-adjusted linear models were then fitted to these network-mean scores to test the same diagnostic contrasts used in the parcel-wise analyses. Resulting p-values were corrected for multiple comparisons using the FDR procedure jointly across both parcels/networks and diagnostic group contrasts.

### Subcortical Gradient Projections

To assess whether the cortical gradient reorganization identified in the primary analyses extended to subcortical circuitry, we computed connectivity-weighted gradient projections for six bilateral subcortical structures and hippocampi, adapting the manifold-projection approach introduced by Park et al. (2021)^59^. Full functional connectivity matrices of dimension 414 × 414 were constructed for each participant, comprising 400 cortical parcels (Schaefer atlas) and 12 subcortical regions plus the hippocampus defined by the Automated Anatomical Labeling (AAL) atlas^60^, encompassing bilateral thalamus, caudate, putamen, pallidum, amygdala, and nucleus accumbens. For each subcortical region i and each gradient G, a weighted gradient projection score was derived as:

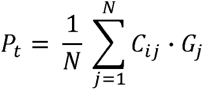

where C_ij_ denotes the functional connectivity between subcortical region i and cortical parcel j, G_j_ is the gradient score of cortical parcel j, and N = 400. This scalar quantity indexes the effective position of each subcortical region within the cortical gradient space, weighted by the strength of its cortical coupling. Left and right homologues were averaged to yield seven bilateral gradient projection scores per participant per gradient. Group differences in subcortical gradient projection scores were tested after residualizing for age and sex using the same one-way ANOVA and FDR-corrected pairwise Welch’s t-test framework applied in the primary cortical analyses, with diagnostic contrasts (HC vs. SZ, HC vs. BD, HC vs. combined patients, SZ vs. BD) evaluated separately for G1 and G2.

### Multivariate Brain–Behaviour Partial Least Squares Regression

To elucidate dimensional brain-behaviour associations across the SZ-BD spectrum, we employed Partial Least Squares (PLS) regression to identify latent dimensions maximizing covariance between brain and behaviour matrices^61–63^. PLS was applied separately for each gradient (G1, G2) to allow gradient-specific brain-behaviour axes to emerge independently. For each gradient, the first latent dimension was retained, indexing the dominant axis of shared brain-behaviour covariance in the combined patient sample (N=77; 37 SZ, 40 BD).

Given the modest sample size, our aim was to find the most generalizable model, which was evaluated through held-out test-set correlations rather than in-sample fit alone during the half-splitting iterations. The brain matrix X (77 × 400) consisted of parcel-wise individual gradient scores, so that PLS operated on aligned cortical-gradient maps. The behavioral matrix Y (77 × 11) comprised diagnosis (schizophrenia, bipolar), symptom dimensions (positive symptoms, disorganization, amotivation, diminished expression, HAMD-21, YMRS), and cognitive composites (attention/working memory, verbal learning, reasoning, vocabulary). Age and sex were first regressed out of the brain matrix across the full patient sample. Feature standardization was then applied using training-partition first: a StandardScaler function was fit exclusively on the training partition for both X and Y, and its parameters (transform function) were subsequently used to standardize the held-out test partition. PLS was then fit on the training partition for each gradient, yielding brain saliences (u) and behavioral saliences (v) defining the first latent dimension. Brain and behavioral scores were computed by projecting the data onto these salience vectors, with test-set generalization assessed via Pearson correlations between these scores. To find the generalizable parameters, 100 random train-test half-splits were evaluated per gradient; the partition minimizing overfitting while maximining test correlations was retained, i.e., maximal performance in the test partition without exceeding training performance, with gradient-specific random seeds reported for full reproducibility^27,48,64^. To facilitate interpretation, brain latent scores were subsequently correlated with each standardized behavioral variable across participants. Statistical significance of these variable-wise associations was assessed using permutation-based Pearson correlations with 1,000 random shuffles and resulting p-values were corrected for multiple comparisons using FDR, separately for each gradient.

### Spatial Correspondence Analysis

To characterize the spatial relationship between the clinical brain–behaviour covariance pattern (PLS brain salience map, LV1) and the normative gradient hierarchy (HC group-mean gradient map), we conducted two complementary analyses at the parcel level (400 Schaefer regions). First, for descriptive visualization, parcel-wise salience weights were plotted against normative gradient values (**Figure 3B**) and summarized by the slope of the ordinary least-squares regression to illustrate the directionality of the clinical signal relative to the normative axis, that is, whether parcels with stronger gradient positions carry proportionally stronger or weaker salience weights. This representation captures the directionality and spatial patterning of the relationship but does not constitute a formal test of whole-brain spatial correspondence, as it does not account for the spatial autocorrelation inherent in cortical maps. To formally test this correspondence, we computed the Pearson correlation between each PLS brain salience map and the HC-group-level gradient map and assessed statistical significance via spin permutation (1,000 permutations), which rotates parcel labels on the cortical surface to preserve spatial autocorrelation while disrupting the correspondence between maps^65,66^. Network-specific contributions are reported in the **Supplementary Results**.

### Subcortical Brain–Behaviour Partial Least Squares

As a complementary extension of the primary cortical PLS, we applied the same partial least squares regression framework (see *Multivariate Brain–Behaviour Partial Least Squares*) to the seven bilateral subcortical gradient projection scores, constituting the brain matrix X (*N* = 77 × 7), paired with the same behavioral matrix Y (*N* = 77 × 11). Subcortical gradient projection values were revisualized for age and sex prior to analysis, with feature standardization and model selection following the identical procedure described above.

### Gradient Similarity Indices

To derive an individual-level representation of the two PLS-defined brain-behaviour patterns, we computed a PLS-based similarity index (SI-PLS) per participant and per gradient. Building on the two PLS analyses described above, SI-PLS projects each individual into a two-dimensional space defined by the G1 and G2 brain salience maps (G1-LV1 and G2-LV1), summarizing in a single scalar per gradient the alignment between a participant’s cortical-gradient topography and the group-derived brain pattern. For each participant and each gradient, SI-PLS was computed as the Pearson correlation between their parcel-wise gradient scores (after age and sex revisualization) and the corresponding PLS brain salience map (u vector from LV1)^67^. Plotting SI-PLS(G1) against SI-PLS(G2) defines a continuous two-dimensional SI-PLS gradient space (G1 × G2) in which each participant occupies a single position. The SI-PLS analysis serves two complementary objectives. First, it provides an integrative individual-level visualization of the PLS findings, projecting each patient onto both PLS dimensions simultaneously; because the salience vectors were derived from the same patient sample, this first objective is explicitly descriptive rather than inferential, and the SZ-BD separation it reveals, reflects the structure already captured by the PLS decomposition. Second, and critically, because healthy controls did not contribute to the PLS fitting, SI-PLS values derived in this group constitute an independent normative reference against which the SZ-BD dimensional pattern was contrasted. Group comparisons involving HC therefore provide externally validated inferences about how the patient-derived gradient dimensions relate to normative cortical organization. Group differences in SI-PLS were tested using the one-way ANOVA and FDR-corrected pairwise Welch’s t-test framework applied in the demographic analyses. As validation, SI-PLS scores were correlated with symptom and cognitive composites within the patient sample (FDR-corrected) to verify that individual similarity indices recapitulate the brain-behaviour structure identified by the PLS.

## Results

### Sample Characteristics

Demographic and clinical characteristics are summarized in **Table 1**. Groups differed significantly in age (F = 4.4, p_FDR_ = 0.01), driven by individuals with SZ being older than HC (p_FDR_ = 0.04), while HC and BD did not differ (p_FDR_ = 0.1). The proportion of male participants also differed significantly across groups (χ² = 8.1, p_FDR_ = 0.02), with a higher proportion of males in the SZ group relative to HC (p_FDR_ = 0.01); sex distribution was comparable between HC and BD, and between SZ and BD. Age and sex were therefore regressed out of all neuroimaging measures. Within the patient sample, individuals with SZ exhibited significantly greater positive symptom severity (t = 7.5, p_FDR_ < 0.001), disorganization (t = 2.1, p_FDR_ = 0.04), diminished expression (t = 3.4, p_FDR_ = 0.002), and amotivation (t = 2.4, p_FDR_ = 0.02) relative to individuals with BD. Depressive and manic symptom burden did not differ significantly between patient groups (HAMD-21: t = −0.7, p = 0.5; YMRS: t = −0.9, p = 0.4). Cognitive impairment was evident across all assessed domains in SZ relative to HC (all p_FDR_ < 0.001). In BD, impairment relative to HC was significant for attention/working memory (p_FDR_ = 0.03) and verbal learning (p_FDR_ < 0.001), but not for reasoning (p_FDR_ = 0.2) or vocabulary (p_FDR_ = 0.7), indicating a more selective cognitive profile. Direct SZ vs. BD comparisons confirmed significantly greater impairment in SZ across all four domains (attention/working memory: p_FDR_ = 0.002; verbal learning: p_FDR_ = 0.001; reasoning: p_FDR_ < 0.001; vocabulary: p_FDR_ < 0.001), with the largest effects observed for verbal learning and vocabulary.

### Global Gradient Architecture and Procrustes Disparity

To establish the normative gradient landscape and evaluate categorical case-control effects on global functional organization, we first characterized the principal connectivity gradients across HC, SZ, and BD groups. The first principal gradient (G1) delineated a continuous sensory axis anchored at one extreme by visual cortices and at the other by somatomotor regions, consistent with the canonical unimodal sensory hierarchy^34,68^. It accounted for 19.4% of the total variance in functional connectivity (**Figure 1A**). The second gradient (G2) captured a broader hierarchical axis, transitioning from primary sensory and motor regions toward higher-order transmodal cortices, most prominently the default mode network, and accounted for an additional 18.3% of the variance. Notably, this unimodal-to-transmodal axis corresponds to what is commonly reported as the first principal gradient^34,68^, with the ordering here reflecting the specific variance structure of the present dataset. Together, G1 and G2 explained 37.7% of the total variance, with a marked drop-off in eigenvalues from the third component onward, supporting their retention as the principal axes for downstream analyses. When projected into the two-dimensional G1-G2 state-space, cortical parcels formed the characteristic triangular manifold, anchored by the visual, somatomotor, and default mode networks, and this global architecture was broadly preserved across HC, SZ, and BD groups (**Figure 1**). For G1, both clinical groups showed significantly greater disparity (D_proc_) relative to HC (HC: 0.32 ± 0.09; SZ: 0.41 ± 0.13, p_FDR_ < 0.001; BD: 0.38 ± 0.13, p_FDR_ = 0.01), indicating that patients with schizophrenia and bipolar disorder deviate more from the normative primary gradient architecture. A similar pattern emerged for G2, with elevated D_proc_ in SZ (0.36 ± 0.09, p_FDR_ < 0.001) and BD (0.33 ± 0.09, p_FDR_ = 0.04) relative to HC (0.29 ± 0.08).

**Figure 1.**
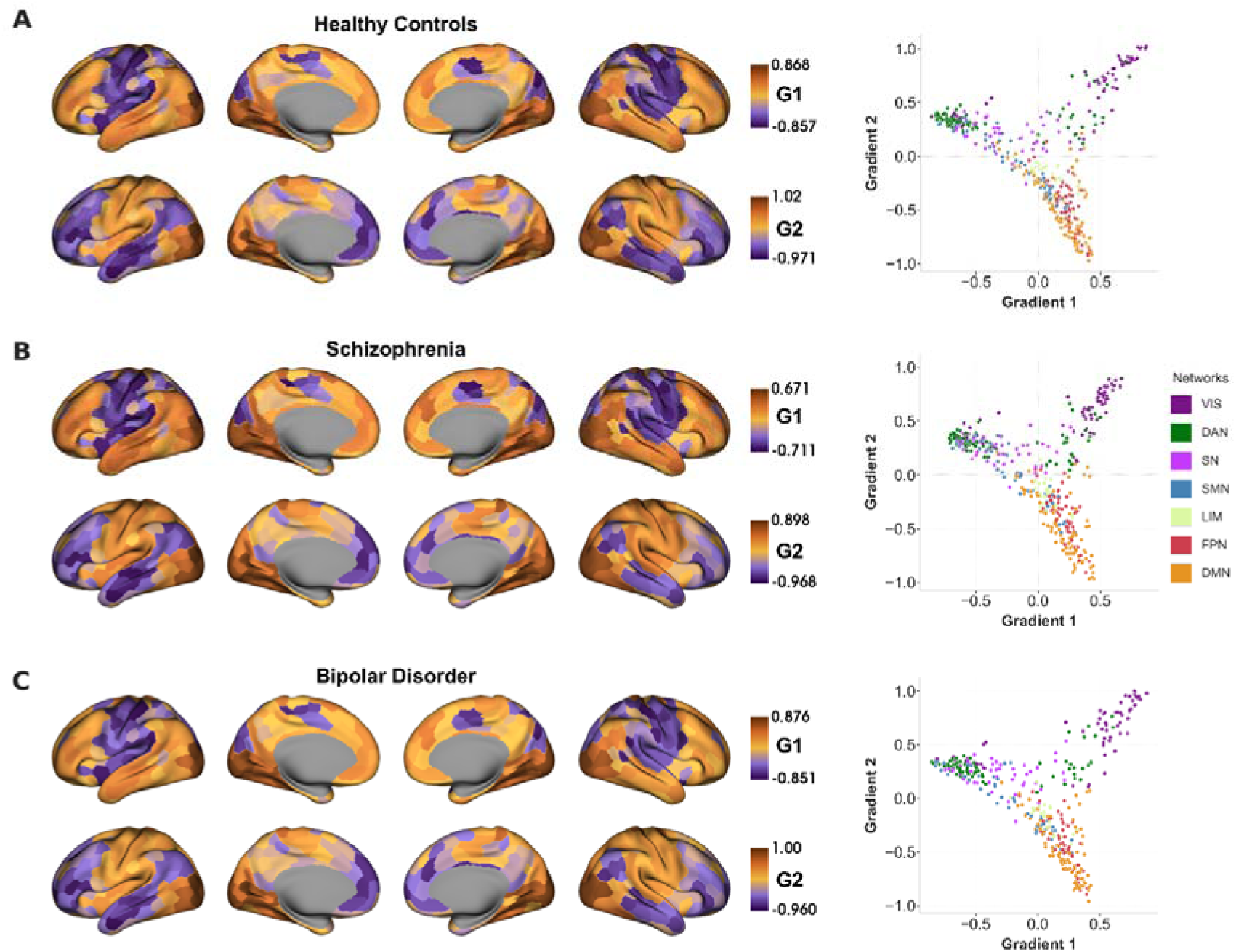
Group-level macroscale cortical functional gradients in healthy controls, schizophrenia, and bipolar disorder. **A.** Healthy controls (HC). Left: surface renderings of the group-mean first (G1; top row) and second (G2; bottom row) cortical gradients on the lateral and medial aspects of both hemispheres; color bars denote gradient-score ranges. Right: two-dimensional gradient space defined by G1 (x-axis) and G2 (y-axis); each point represents one Schaefer parcel, colored according to its assignment to one of seven canonical resting-state networks. **B.** Schizophrenia (SZ) and **C.** Bipolar disorder (BD) are displayed with the same layout and conventions. Abbreviations: VIS, visual; DAN, dorsal attention; SN, salience; SMN, somatomotor; LIM, limbic; FPN, frontoparietal; DMN, default mode network.

### Network-Level Topographic Alterations

To localize categorical case-control effects on macroscale functional topography, we fit linear models at both the parcel and network levels. Parcel-wise tests yielded no parcels surviving FDR correction in any group comparison (HC vs. SZ, HC vs. BD, HC vs. combined patients). At the network level, however, linear models revealed selective and diagnostically patterned deviations in functional topography despite preservation of the global manifold structure (**Figure 2A**). Along both gradient axes, the frontoparietal network emerged as the most consistently affected system. Along G1, the combined patient group scored significantly lower than HC (t = -1.99, p_FDR_ = 0.04), reflecting a broad compression of the frontoparietal position along the primary sensory axis; this transdiagnostic effect was accompanied by a disorder-specific reduction in visual network scores in SZ alone (t = -2.05, p_FDR_ = 0.04), indicating a relative contraction of the visual anchor of the sensory hierarchy. Along G2, frontoparietal elevation was the most robustly observed effect, present in SZ (t = 2.61, p_FDR_ = 0.01), in BD (t = 2.23, p_FDR_ = 0.03), and most strongly in the combined patient group (t = 3.09, p_FDR_ < 0.01), reflecting a transdiagnostic compression away from the normative transmodal position. The somatomotor network showed a complementary disorder-specific pattern, with reduced G2 scores in BD (t = −2.29, p_FDR_ = 0.02) and in the combined patient sample (t = −2.44, p_FDR_ = 0.02), indicating a relative displacement of the somatomotor anchor away from the unimodal pole of the G2 axis. Statistical results for all network comparisons are provided in the **Supplementary Results** (**Supplementary Results, Table S1**).

**Figure 2.**
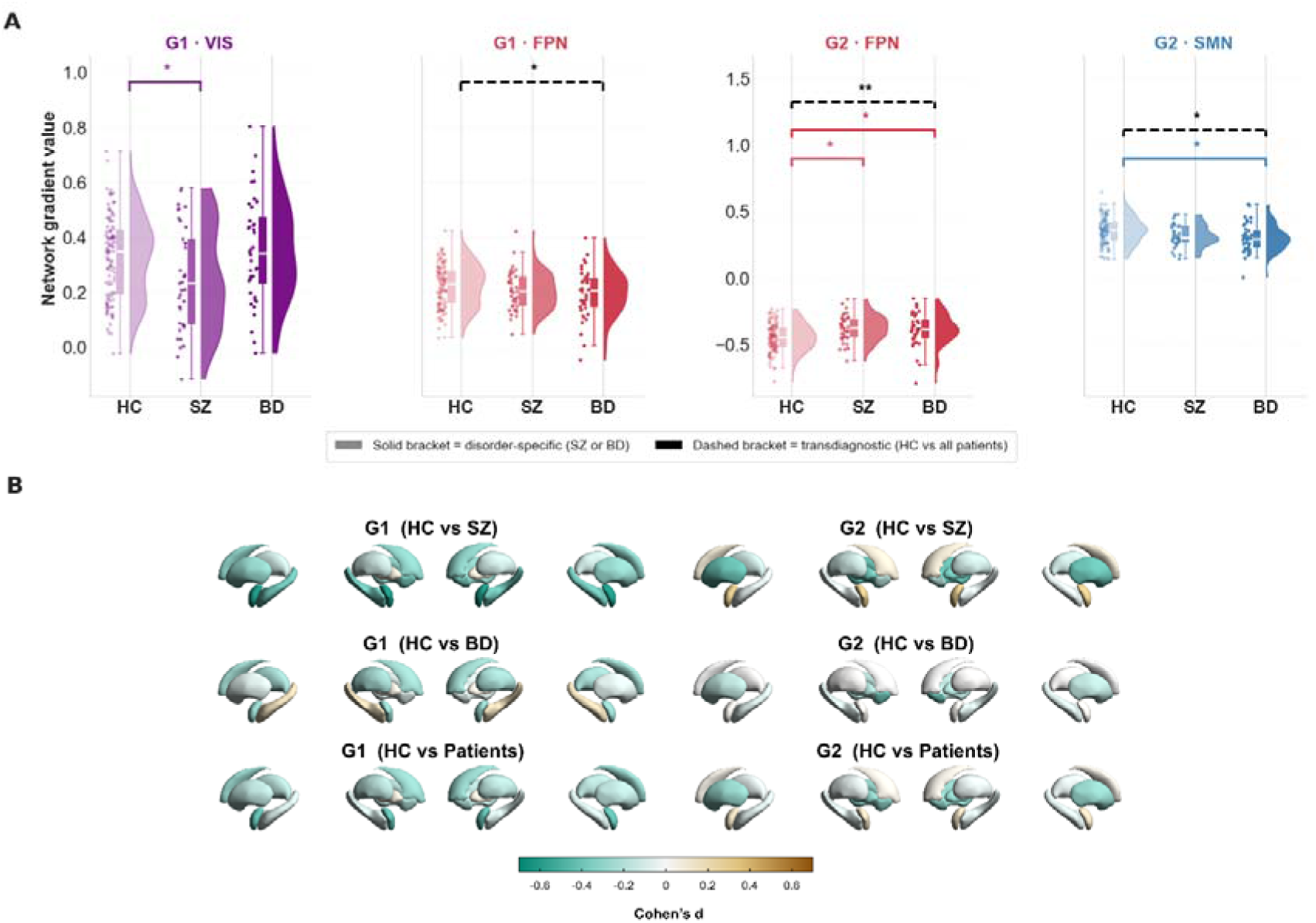
Network-level and subcortical gradient projection alterations across diagnostic groups. **A.** Raincloud plots depicting individual network-mean gradient values for all four significant network-gradient combinations. Solid brackets denote disorder-specific comparisons (HC vs. SZ or HC vs. BD); dashed brackets denote transdiagnostic comparisons (HC vs. combined patients). *p < 0.05, **p < 0.01 (FDR-corrected). **B.** Cohen’s d effect sizes for G1 (left column) and G2 (right column) gradient projections onto bilateral subcortical structures and hippocampi. Three contrasts are shown (top to bottom): HC vs. SZ, HC vs. BD, and HC vs. all patients combined. Negative values (green) indicate lower projection scores in patients relative to controls; positive values (yellow) indicate higher scores. Abbreviations: VIS, Visual; FPN, Frontoparietal; SMN, Somatomotor; HC, Healthy Controls; SZ, Schizophrenia; BD, Bipolar Disorder.

### Subcortical Gradient Projection Alterations

To test whether the cortical gradient reorganization identified above extended to subcortical circuitry, we examined group differences in connectivity-weighted gradient projection scores across seven bilateral subcortical structures (full per-region statistics in **Supplementary Table S2**). Along G1, a single FDR-significant effect emerged: SZ individuals showed a lower amygdala gradient projection score relative to HC (t = −3.14, d = −0.60, p_FDR_ = 0.03), indicating a reduced effective coupling of the amygdala along the sensory hierarchy axis; this effect was not significant in the combined patient group. No further subcortical regions reached FDR-corrected significance under any contrast. Effect sizes for all bilateral subcortical structures across the three contrasts are visualized in **Figure 2B**.

### Cortical Brain–Behaviour Associations (PLS)

To identify dimensional brain-behaviour associations across the SZ-BD spectrum, we applied PLS to the combined patient sample, linking parcel-wise gradient scores to diagnostic, symptom, and cognitive measures (**Figure 3**). For G1, the brain-behaviour correlation in the first latent dimension (LV1) was significant (training *r* = 0.65, test *r* = 0.48; **Supplementary Results, Figure S1A**). The corresponding behavioral salience pattern was driven primarily by bipolar disorder diagnosis and by lower positive symptom severity, disorganization, and negative symptom burden in BD relative to SZ, consistent with the comparatively milder clinical phenotype observed in BD relative to SZ (**Figure 3C**). On the brain side, G1-LV1 exhibited a distributed cortical topography encompassing sensory and association networks (**Figure 3A**). For G2, the single latent dimension also yielded a significant brain-behaviour correlation (train *r* = 0.74, test *r* = 0.41; **Supplementary Results, Figure S1A**). In contrast to G1, the G2-LV1 behavioral salience pattern was characterized by schizophrenia diagnosis, greater positive and negative symptom severity, and poorer cognitive performance across all assessed domains, indicating that this dimension captured a more severe and cognitively impaired clinical profile (**Figure 3C**). The corresponding brain salience map highlighted complementary shifts in large-scale network organization along G2 (**Figure 3A**). The two latent dimensions thus yielded spatially dissociable cortical salience maps, consistent with the recovery of two distinct gradient-specific brain-behaviour axes rather than redundant projections of a shared behavioral latent structure; the approximately balanced diagnostic composition of the sample (SZ, N = 37; BD, N = 40) further argues against this dissociation arising from asymmetric group contributions to the PLS decomposition. Finally, we repeated our brain-behaviour PLS using subcortical gradient projection scores; both latent dimensions showed markedly attenuated out-of-sample generalization relative to the cortical PLS, indicating that the detectable brain-behaviour covariance was predominantly supported by cortical gradient organization (see **Supplementary Results, Figure S1B**).

**Figure 3.**
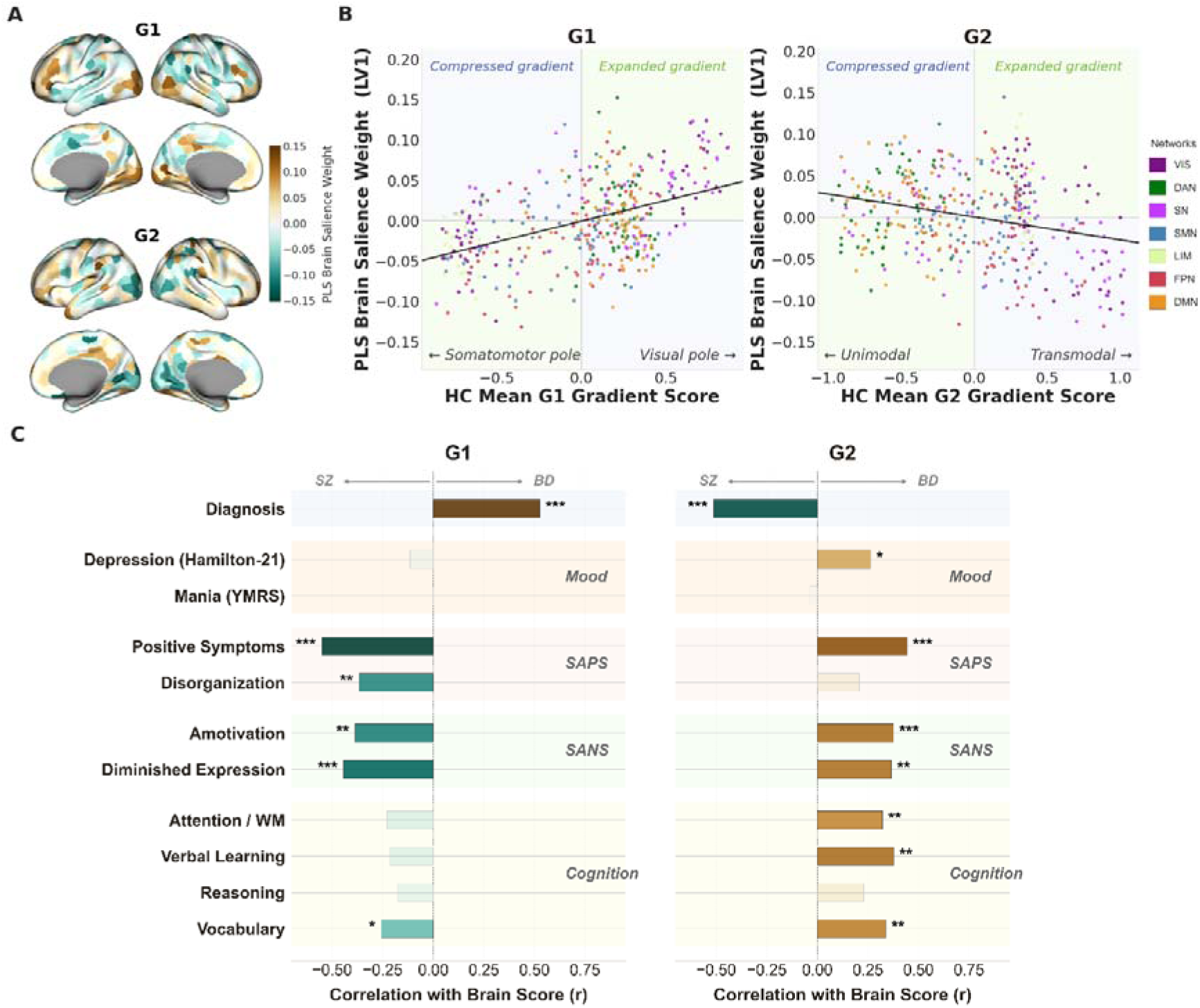
Associations between cortical gradients and clinical phenotypes. **A.** Brain salience maps for the first latent variable (LV1) derived separately for Gradient 1 (G1) and Gradient 2 (G2). Warm and cool colors indicate positive and negative brain salience weights. **B.** Relationship between normative gradient position (HC group mean gradient score) and PLS brain salience weight (LV1) across the 400 parcels, colored by functional network. A positive slope (G1-LV1, r = 0.450, p < .001) indicates that the clinical brain pattern expands the G1 axis; a negative slope (G2-LV1, r = −0.267, p < .001) indicates compression of the G2 axis. Green and blue quadrants denote gradient-expanded and gradient-compressed regions, respectively. **C.** Correlations between brain latent scores and behavioral variables for G1-LV1 and G2-LV1. Asterisks indicate FDR-corrected significance, * p_FDR_ < 0.05; ** p_FDR_< 0.01; *** p_FDR_< 0.001. Abbreviations: BD, Bipolar Disorder; DAN, Dorsal Attention Network; DMN, Default Mode Network; FPN, Frontoparietal Network; LIM, Limbic; LV, Latent Variable; SANS, Scale for the Assessment of Negative Symptoms; SAPS, Scale for the Assessment of Positive Symptoms; SMN, Somatomotor Network; SN, Salience Network; SZ, Schizophrenia; VIS, Visual Network; WM, Working Memory; YMRS, Young Mania Rating Scale.

### Spatial Correspondence of PLS Brain Salience Maps

Spatial correspondence between each PLS brain salience map (LV1) and the HC group-mean gradient map was assessed using two complementary parcel-wise analyses (**Figure 3B**). Descriptively, the scatter of parcel-wise salience weights against normative gradient values showed a positive slope for G1-LV1 (β = +0.45), indicating that parcels positioned further along the G1 axis carried proportionally stronger salience weights in the same direction, consistent with an expansion of the sensory poles relative to HC. This is mirrored by the disorder-specific visual pole compression identified for SZ in the categorical analyses: whereas SZ showed contraction of the G1 sensory axis, the BD-associated PLS pattern runs in the opposite direction, reflecting a relatively preserved sensorimotor-to-visual differentiation that aligns with lower symptom burden. For G2-LV1, the slope was negative (β = −0.27), indicating that the SZ-associated signal ran counter to the normative G2 axis, with frontoparietal parcels as the primary driver. To formally test whole-brain spatial correspondence while accounting for spatial autocorrelation, spin permutation tests (1,000 permutations) were applied: no significant correspondence emerged for G1 (r = +0.07, p_spin_ = 0.20), whereas G2 showed a significant negative correspondence (r = −0.23, p_spin_ < 0.001). Together, these analyses indicate that the G1 brain salience map is descriptively congruent with the normative G1 axis but does not survive correction for spatial autocorrelation, whereas the G2 brain salience map runs counter to the normative transmodal hierarchy at both descriptive and formal levels. Network-specific contributions are reported in the **Supplementary Results**.

### Individual-Level Gradient Similarity Indices (SI-PLS)

To translate group-level gradient reorganization patterns into individual-level measures, we computed two PLS-derived similarity indices, SI-PLS(G1) and SI-PLS(G2), each capturing how strongly a given participant’s cortical gradient topography aligns with the corresponding PLS brain salience map (G1-LV1 and G2-LV1, respectively). Together, these two scalar indices define a joint SI-PLS gradient space (G1×G2): a continuous two-dimensional similarity space in which each participant occupies a single position, enabling subject-level localization along the SZ-BD spectrum independently of categorical diagnosis. This two-dimensional representation provides the most informative visualization of the individual-level pattern and is shown in **Figure 4A**. Group comparisons were then used to test whether the dimensional PLS pattern derived from the combined patient sample differed from the healthy control cohort, which had not contributed to the PLS fitting. Both SI-PLS indices showed significant diagnostic effects (all ANOVA p < .001; **Figure 4B**). Given that the PLS salience vectors were derived from the combined patient sample, the SZ–BD separation in SI-PLS space is best interpreted as an individual-level re-expression of the PLS decomposition rather than an independent replication; the comparisons involving HC are the inferentially novel findings. For SI-PLS along G1 (F = 12.08, p < 0.001), individuals with BD showed significantly higher scores than both HC (t = −2.53, p_FDR_ = 0.014) and SZ (t = −4.59, p_FDR_ < 0.001), while SZ scored significantly lower than HC (t = 3.21, p_FDR_ = 0.003). This pattern indicates that BD patients most strongly express the G1 brain salience pattern, whereas SZ patients show the least alignment with it, consistent with G1-LV1 indexing a bipolar-associated gradient reorganization. For SI-PLS along G2 (F = 15.84, p_FDR_ < 0.001), SZ showed significantly elevated scores relative to both HC (t = −5.17, p_FDR_ < 0.001) and BD (t = 4.92, p_FDR_ < 0.001), whereas HC and BD did not differ (p_FDR_ = 0.2), consistent with G2-LV1 capturing a schizophrenia-associated reorganization pattern. Notably, the confidence ellipses in Figure 4A revealed substantial within-group spread and partial overlap between diagnostic groups, indicating that individual position within the joint SI-PLS space was not fully determined by diagnostic category. Validation analyses confirmed that SI-PLS scores were associated with symptoms and cognitive measures within the patient sample (**Supplementary Results, Figure S3**).

**Figure 4.**
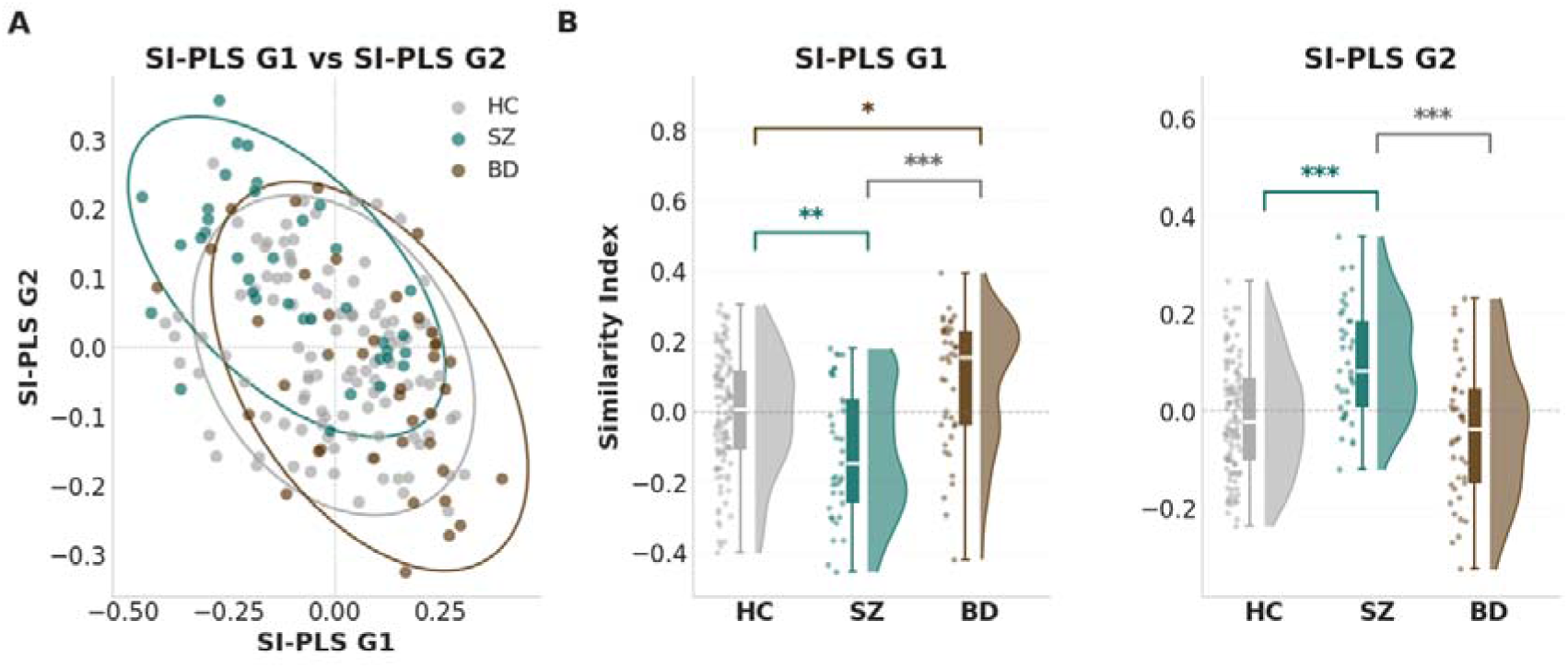
Individual expression of group-level gradient reorganization patterns. **A.** Two-dimensional scatter plot of SI-PLS G1 versus SI-PLS G2, with 95% confidence ellipses per diagnostic group. **B.** Raincloud plots depicting group differences in PLS derived similarity indices (SI-PLS) along G1 and G2 across diagnostic groups. Colored brackets denote disorder-specific comparisons (HC vs. SZ or HC vs. BD); grey brackets denote transdiagnostic comparisons (HC vs. combined patients). Abbreviations: HC, Healthy Controls; SZ, Schizophrenia; BD, Bipolar Disorder.

## Discussion

In this study, we characterized macroscale cortical gradient organization across the schizophrenia-bipolar spectrum, examining how functional brain architecture differs between diagnostic groups and varies continuously at the individual level. Adopting an integrative framework including categorical case-control, multivariate dimensional, and individual-level similarity index analyses, three key findings emerged. First, while the global triangular manifold anchored by the visual, somatomotor, and default-mode networks was broadly preserved in SZ and BD, group-level case-control comparisons revealed systematic deviations from the healthy template along both the visual-to-somatomotor (G1) and unimodal-to-transmodal (G2) axes in both disorders. Second, multivariate PLS revealed contrasting brain-behavior patterns for the two gradients, with G1 expressing a predominantly BD-associated profile of comparatively lower symptom burden and G2 expressing a predominantly SZ-associated profile linked to greater cognitive impairment and psychotic symptom severity. Third, projecting individuals into the joint SI-PLS gradient space (G1 × G2) revealed a graded transdiagnostic separation along both principal axes, with SZ and BD occupying largely non-overlapping regions of this two-dimensional space and HC positioned between them.

The first key observation is that the global triangular manifold remained recognizable across groups, while Procrustes disparity along both G1 and G2 was significantly elevated in patients. This pattern suggests that group-level deviations along both principal gradient axes, rather than global topographic disorganization, distinguish SZ and BD from controls. This pattern is reminiscent of earlier observations in autism, where the gradient hierarchy is preserved but subject-specific precision is reduced^38^ and finds a more direct parallel in recent psychosis work documenting the same dissociation^46^. It is further consistent with reports of substantial interindividual heterogeneity in functional brain organization across the psychosis spectrum^2^. The graded rather than categorical nature of this deviation across SZ and BD is compatible with a dimensional framing of differences in psychosis^20^.

Parallel to this globally preserved architecture, network-level analyses revealed a mixed pattern of disorder-specific and transdiagnostic effects. On the visual-to-somatomotor axis, the visual pole was specifically compressed in schizophrenia but not in bipolar disorder. Using a different gradient construction approach in a partially overlapping SZ cohort, this finding corroborates the schizophrenia-specific secondary-gradient disruption reported by Holmes et al. (2023)^46^, supporting the robustness of this visual-network signature across methodological choices^69^, and extends it by demonstrating that it is specific to schizophrenia across the SZ-BD spectrum. On the unimodal-to-transmodal axis, whereas prior work documented somatomotor compression in schizophrenia^43,44^, in our transdiagnostic sample this effect was significant only in bipolar disorder. This divergence may reflect clinical heterogeneity within schizophrenia, where a subgroup with greater mood symptom burden drives somatomotor involvement, a pattern that becomes more apparent when bipolar disorder is included in the comparison. Across both axes, the frontoparietal network emerged as the most consistent transdiagnostic finding, showing compression in both patient groups, confirming and extending the frontoparietal disruption reported by Dong et al. (2023)^43^ in schizophrenia to the broader SZ-BD spectrum, consistent with meta-analytic evidence of frontoparietal involvement in schizophrenia^70^, bipolar disorder^71^, and the psychosis spectrum more broadly^72^. More generally, these findings reveal a dissociation between the two principal gradient axes: the unimodal-to-transmodal axis carries predominantly transdiagnostic alterations centered on higher-order frontoparietal systems, whereas the visual-to-somatomotor axis captures more disorder-specific effects, with visual compression selective to schizophrenia and somatomotor displacement more prominent in bipolar disorder.

A complementary picture emerged from subcortical-cortical connectivity analysis, in which we used connectivity-weighted gradient projections to assess how each subcortical region is effectively situated within the cortical gradient space. Among the seven bilateral structures examined, the amygdala emerged as the principal locus of subcortical-cortical gradient involvement: SZ participants showed a lower G1 score relative to HC, indicating that the amygdala’s effective alignment with the sensorimotor-to-visual axis was shifted in SZ. This pattern fits naturally with the established role of amygdala dysconnectivity in the affective and salience-processing disturbances of schizophrenia^73–75^, and parallels reports of disrupted subcortical functional hierarchy in first-episode illness^76^. The amygdala thus provides a subcortical anchor for the SZ-related signal carried by G1, consistent with cortical gradient reorganization extending to specific subcortical regions through their cortical connectivity.

Whereas categorical comparisons localized network-level disruptions independent of clinical heterogeneity, multivariate PLS revealed which patterns of gradient reorganization co-vary with the dimensional clinical-cognitive profile of patients. Along G1, bipolar disorder was associated with a relatively preserved and descriptively expanded gradient architecture alongside comparatively lower symptom burden, contrasting with the visual pole compression identified for schizophrenia in the categorical comparisons. Along G2, the predominant signal was SZ-associated, mirroring the transdiagnostic frontoparietal compression identified in the case-control analyses but now revealing its dimensional clinical correlation: greater positive and negative symptom severity and broad cognitive impairment. The two patterns also differed in how they aligned with the normative gradient architecture. The BD-associated G1 signal was positively aligned with the normative G1 axis, consistent with an expansion of the sensorimotor-to-visual differentiation, with the visual and somatomotor poles diverging further relative to HC. The SZ-associated G2 signal, by contrast, was negatively aligned with the normative G2 axis, with frontoparietal regions as the primary driver, indicating a reduced separation between unimodal and transmodal systems. Thus, G1 and G2 did not simply carry different clinical content; they did so in opposing relations to the normative gradient architecture. This dissociation is consistent with the view that G1 and G2 represent two functionally distinct axes of cortical organization^34,68^ and suggests that clinical variance across the psychosis spectrum is distributed differently across these two axes rather than converging on a single dimension of pathology.

At the individual level, these gradient-specific effects cohered into a clinically informative geometry. Using PLS-derived similarity indices (SI-PLS), scalar measures that capture how strongly each participant’s cortical gradient topography aligns with the PLS-derived brain salience pattern, placing individuals continuously within the gradient–behaviour space, we show that diagnostic groups occupied largely non-overlapping regions of the gradient-behaviour coupling space. BD participants cluster toward positive SI-PLS G1 and near-zero SI-PLS G2, SZ toward positive SI-PLS G2, and HC between them. Because HC were withheld from the PLS estimation, their intermediate positioning in the joint SI-PLS space constitutes the principal inferentially independent finding of this analysis: the gradient dimensions that dissociate SZ from BD are organized along a continuum on which healthy controls occupy an intermediate, rather than categorically separate, position. The separation was graded rather than absolute, with individuals distributed continuously across the space such that diagnostic categories organized its overall structure without fully determining individual position. The clinical signature of each disorder was therefore not a single scalar deviation along a common axis of pathology nor a discrete categorical location, but a position in a two-dimensional neural space along which diagnostic groups showed clustering with meaningful individual-level spread. In this sense, the two disorders occupied distinct but partially overlapping regions of a shared neural substrate, a configuration naturally compatible with dimensional accounts of psychosis in which SZ and BD share common neural axes while expressing them in different combinations and degrees^10,12,13^, and one in which individual position, rather than group membership alone, may be the more informative descriptor of where a given participant sits within the psychosis spectrum.

## Limitations

Several limitations should be noted. First, the sample, though transdiagnostic, is modest in size, limiting statistical power for fine-grained subgroup analyses and the detection of weaker effects, particularly in subcortical gradient projection analyses. Replication in larger, independent cohorts is necessary to establish the generalizability of the gradient similarity indices and their clinical correlates. Second, the cross-sectional design precludes inference about the effects of illness course or antipsychotic treatment. In a partially overlapping cross-sectional sample, medication was not associated with cortical gradients^46^; more recently, however, subcortical limbic gradients were shown to change over twelve months of treatment in first-episode psychosis^75^, suggesting potential modest normalizing effects at the subcortical level. The individual gradient-based similarity indices introduced here provide a foundation for future longitudinal investigations in larger samples to evaluate their predictive utility for illness course, treatment response, and medication effects, questions with direct clinical relevance. Third, individuals with SZ in this sample were older and more predominantly male; although age and sex were regressed from all neuroimaging measures, residual confounding cannot be fully excluded.

## Conclusions

Taken together, these findings describe a cortical gradient landscape structured by two principal axes carrying distinct clinical signals in SZ and BD. Categorical comparisons identified both shared alterations, especially in higher-order frontoparietal systems, and diagnosis-specific effects, including visual contraction in SZ and somatomotor displacement in BD, which are more evident in primary sensory networks. At the dimensional level, G1 co-varied with BD-related variation while G2 co-varied with SZ-related symptom and cognitive burden, and projecting individuals into the joint SI-PLS gradient space revealed a graded two-dimensional separation invisible from either axis alone, a configuration more consistent with a multi-axis dimensional framework than with a strictly categorical view of the SZ-BD spectrum. The gradient similarity indices introduced here illustrate one tractable way to position patients within this space at the subject level which could inform biomarker development of dimensional brain-behaviour phenotypes in future longitudinal studies. Collectively, this work characterizes transdiagnostic alterations in higher-order systems alongside disorder-specific alterations in primary sensory systems and introduces a gradient-based framework for capturing clinical heterogeneity across the SZ-BD spectrum at the individual level.

## Data Availability

All data produced in the present study are available upon reasonable request to the authors

## Author Contributions

<u>Asia Ferrari:</u> Conceptualization, Formal analysis, Data curation, Methodology, Investigation, Software, Visualization, Writing - original draft; <u>Bin Wan:</u> Methodology, Formal analysis, Software, Writing - review & editing; <u>Judith Kabbeck:</u> Conceptualization, Data curation, Investigation, Writing - review & editing; <u>Amin Saberi:</u> Methodology, Software, Writing - review & editing; <u>Stefan Kaiser:</u> Resources, Writing - review & editing; <u>Valeria Kebets:</u> Data curation, Resources, Writing - review & editing; <u>Clara A. Moreau:</u> Writing - review & editing; <u>Paul Thompson:</u> Resources, Writing - review & editing; <u>Theo Van Erp:</u> Resources, Writing - review & editing; <u>Jessica A. Turner:</u> Resources, Writing - review & editing; <u>B.T. Thomas Yeo</u>: Data curation, Resources, Writing - review & editing; <u>Boris C.</u> <u>Bernhardt:</u> Resources, Methodology, Supervision, Writing - review & editing; <u>Sofie Valk:</u> Conceptualization, Methodology, Supervision, Writing - review & editing; <u>Matthias Kirschner:</u> Conceptualization, Methodology, Project administration, Supervision, Writing - original draft.

## Acknowledgements

The authors are particularly grateful to B.T.T. Yeo and V. Kebets for kindly sharing the preprocessed resting-state fMRI data used in this work. B.C.B. acknowledges support from the Canadian Institutes of Health Research (CIHR), National Sciences and Engineering Research Council of Canada

(NSERC), the Centre of Excellence in Epilepsy at the Neuro (CEEN), Canada Research Chairs Program (CRC), and Healthy Brains, Healthy Lives (HBHL).

