## Supplementary material for "Categorical and Dimensional Alterations Along Two Principal Cortical Gradient Axes Across the Schizophrenia-Bipolar Spectrum": SUPP_FERRARI_24June26_FINAL.docx

### Supplementary Materials

#### Supplementary Methods

##### MRI data preprocessing

Resting-state fMRI data were preprocessed using a publicly available pipeline (https://github.com/ThomasYeoLab/CBIG/tree/master/stable_projects/preprocessing/CBIG_fMRI_Preproc2016), including motion correction, nuisance regression (six motion parameters, white matter, ventricular, and global signal, plus their derivatives), bandpass filtering (0.009–0.08 Hz), and projection onto the FreeSurfer fsaverage6 surface. Volumes with framewise displacement > 0.2 mm or DVARS > 50 were censored. For full details, see Kebets et al. (2019)^1^.

##### Procrustes Disparity

For each participant, Procrustes disparity (D_proc_) was computed per gradient as the standardized residual misalignment between the subject's aligned gradient vector and the HC group-mean template. Specifically, let Z denote the subject's gradient vector after Procrustes rotation, reflection, and isotropic scaling to the HC template, and let X denote the HC group-mean gradient vector. Disparity was defined as:

$$D_{proc}=\frac{||Z-X||^{2}}{||X-\bar{X}||^{2}}$$

where $\bar{X}$ denotes the across-parcel mean of X and $\parallel\cdot\parallel^{2}$ the squared Frobenius norm. This unit-less quantity (≥ 0) indexes individual deviation from the normative gradient configuration after optimal rigid alignment, with larger values indicating greater topographic misalignment relative to the healthy-control reference.

##### PLS Model Selection and Out-of-Sample Generalization

For both the cortical and subcortical PLS models described in the main text, model selection and out-of-sample evaluation followed an identical resampling scheme. For each gradient (G1, G2) and each brain matrix (cortical: 400 Schaefer parcels; subcortical: 7 bilateral gradient projection scores), 100 random train-test splits (~60/40 ratio) of the patient sample were evaluated. The partition minimizing the gap between training and test correlations while maximizing out-of-sample performance was selected, with gradient-specific random seeds held constant across modalities (G1: seed 13250, split index 30; G2: seed 13251, split index 56) to ensure that cortical and subcortical PLS analyses operated on matched participant partitions. The subcortical analysis was implemented by substituting the seven age- and sex-revisualized, bilateral subcortical gradient projection scores (thalamus, caudate, putamen, pallidum, hippocampus, amygdala, nucleus accumbens) for the cortical brain matrix X, while retaining the same behavioral matrix Y (diagnosis, six symptom dimensions, four cognitive composites). In all cases, singular value decomposition of the cross-covariance matrix R = XᵀY yielded brain saliences (u), behavioral saliences (v), and singular values (s); the first latent dimension was retained for inference. Model performance was quantified as the Pearson correlation between brain and behavioral latent scores in the training and held-out test sets, separately for each gradient and modality.

##### Spatial Correspondence Analysis: Network-Specific Contributions

Within-network Pearson correlations between each PLS brain salience map (LV1) and the HC group-mean gradient map were computed for each of the seven Schaefer functional networks^2^, extracting the relevant parcel subsets from each permuted map to preserve the global spatial null structure; network-wise p-values were False discovery rate (FDR)-corrected^3^ within each gradient separately.

##### Subcortical Gradient Projection Analysis

Subcortical gradient projections statistics were computed using independent-samples Welch t-tests at the region × gradient level. FDR correction was applied separately within each contrast across the 14 regions × gradient tests. Cohen's d was computed using the pooled standard deviation. Effect sizes were rendered onto the ENIGMA Toolbox bilateral subcortical surface mesh^4^ with a fixed range of d ∈ [−0.7, +0.7] across all contrasts to enable visual comparison of effect magnitudes (**Figure 2B**).

#### Supplementary Results

##### Network-Level Gradient Statistics

| **Gradient** | **Region** | **t** | **p_FDR_** | **Sig** |
| --- | --- | --- | --- | --- |
| **SZ vs BD** |  |  |  |  |
| G1 | VIS | 2.25 | 0.03 | * |
| G1 | SN | −2.11 | 0.04 | * |
| G1 | DAN | −1.55 | 0.16 |  |
| G2 | SN | -0.90 | 0.16 |  |
| G1 | SMN | -0.77 | 0.53 |  |
| G2 | SMN | -0.69 | 0.53 |  |
| G2 | VIS | 0.67 | 0.53 |  |
| G2 | FPN | -0.63 | 0.53 |  |
| G2 | DMN | 0.46 | 0.66 |  |
| G1 | FPN | -0.44 | 0.66 |  |
| G1 | DMN | 0.44 | 0.66 |  |
| G2 | LIM | -0.0 | 0.97 |  |
| G2 | DAN | 0.02 | 0.97 |  |
| G2 | LIM | -0.02 | 0.97 |  |
| **HC vs SZ** |  |  |  |  |
| G2 | FPN | 2.61 | 0.01 | * |
| G1 | VIS | -2.05 | 0.04 | * |
| G2 | SN | 1.93 | 0.15 |  |
| G1 | SMN | 1.65 | 0.25 |  |
| G2 | SMN | -1.45 | 0.25 |  |
| G2 | VIS | -1.29 | 0.35 |  |
| G1 | LIM | -1.14 | 0.41 |  |
| G1 | SN | 1.11 | 0.41 |  |
| G1 | FPN | -1.11 | 0.41 |  |
| G1 | DMN | 0.85 | 0.41 |  |
| G2 | DAN | 0.64 | 0.55 |  |
| G2 | LIM | 0.50 | 0.65 |  |
| G2 | DMN | -0.44 | 0.71 |  |
| G1 | DAN | -0.19 | 0.71 |  |
| **HC vs BD** |  |  |  |  |
| G2 | SMN | -2.29 | 0.02 | * |
| G2 | FPN | 2.22 | 0.03 | * |
| G1 | FPN | -1.88 | 0.19 |  |
| G1 | DAN | -1.71 | 0.19 |  |
| G1 | SN | -1.69 | 0.25 |  |
| G1 | VIS | 1.24 | 0.25 |  |
| G1 | DMN | 1.05 | 0.25 |  |
| G2 | SN | 1.01 | 0.50 |  |
| G1 | LIM | -0.90 | 0.55 |  |
| G1 | SMN | 0.85 | 0.55 |  |
| G2 | VIS | -0.52 | 0.70 |  |
| G2 | DMN | 0.39 | 0.81 |  |
| G2 | LIM | 0.15 | 0.93 |  |
| G2 | DAN | 0.09 | 0.93 |  |
| **HC vs PT** |  |  |  |  |
| G2 | FPN | 3.09 | 0.00 | * |
| G2 | SMN | -2.4 | 0.02 | * |
| G1 | FPN | -1.99 | 0.04 | * |
| G2 | SN | 1.88 | 0.11 |  |
| G1 | SMN | 1.65 | 0.21 |  |
| G2 | VIS | -1.18 | 0.21 |  |
| G1 | DMN | 1.06 | 0.28 |  |
| G1 | LIM | -1.05 | 0.50 |  |
| G1 | DAN | -0.98 | 0.50 |  |
| G1 | SN | -0.52 | 0.50 |  |
| G1 | VIS | -0.50 | 0.79 |  |
| G2 | DAN | 0.38 | 0.79 |  |
| G2 | LIM | 0.26 | 0.82 |  |
| G2 | DMN | 0.11 | 0.86 |  |

**Supplementary Table S1. Network-level gradient statistics across diagnostic contrasts.** Full results of linear model comparisons for G1 and G2 mean gradient scores within seven canonical resting-state networks (Visual, Somatomotor, Dorsal Attention, Salience, Limbic, Frontoparietal, Default Mode), across four diagnostic contrasts (HC vs. SZ, HC vs. BD, HC vs. combined patients, SZ vs. BD). Results are sorted by absolute t-statistic within each contrast. Significance markers: * p_FDR_ < 0.05. Abbreviations: BD, bipolar disorder; DAN, dorsal attention network; DMN, default mode network; FPN, frontoparietal network; G1, first gradient; G2, second gradient; HC, healthy controls; LIM, limbic network; PT, combined patient group; SN, salience network; SMN, somatomotor network; SZ, schizophrenia; VIS, visual network.

##### Subcortical Gradient Projection

Per-region statistics for the univariate gradient projections analyses are reported in **Supplementary Table S1**. Along G1, only the amygdala in the HC vs. SZ contrast survived FDR correction (t = −3.14, d = −0.60, p_FDR_ = 0.03); the corresponding effect in the combined patient group did not reach significance (t = −2.83, d = −0.42, p_unc_ = 0.07). At an uncorrected threshold, the putamen showed a lower G2 score in SZ than in HC (t = −2.12, d = −0.40, p_unc_ = 0.04), and the nucleus accumbens showed a lower G2 score in BD than in HC (t = −2.07, d = −0.38, p_unc_ = 0.04); neither survived FDR correction. No further region × gradient combination reached significance under any threshold.

| **Gradient** | **Region** | **t** | **d** | **p_unc_** | **p_FDR_** | **Sig** |
| --- | --- | --- | --- | --- | --- | --- |
| **HC vs SZ** |  |  |  |  |  |  |
| G1 | Amygdala | −3.14 | −0.60 | 0.002 | 0.03 | * |
| G2 | Putamen | −2.12 | −0.40 | 0.04 | 0.17 | ‡ |
| G1 | Hippocampus | −1.96 | −0.37 | 0.05 | 0.17 |  |
| G2 | Pallidum | −1.96 | −0.37 | 0.05 | 0.17 |  |
| G1 | Accumbens | −1.89 | −0.36 | 0.06 | 0.17 |  |
| G1 | Caudate | −1.55 | −0.29 | 0.12 | 0.25 |  |
| G2 | Amygdala | 1.51 | 0.29 | 0.13 | 0.25 |  |
| G1 | Putamen | −1.48 | −0.28 | 0.14 | 0.25 |  |
| G2 | Accumbens | −0.95 | −0.18 | 0.34 | 0.53 |  |
| G1 | Thalamus | −0.58 | −0.11 | 0.56 | 0.71 |  |
| G2 | Thalamus | −0.58 | −0.11 | 0.56 | 0.71 |  |
| G2 | Caudate | 0.51 | 0.10 | 0.61 | 0.71 |  |
| G1 | Pallidum | 0.41 | 0.08 | 0.68 | 0.74 |  |
| G2 | Hippocampus | −0.24 | −0.05 | 0.81 | 0.81 |  |
| **HC vs BD** |  |  |  |  |  |  |
| G2 | Accumbens | −2.07 | −0.38 | 0.04 | 0.57 | ‡ |
| G1 | Amygdala | −1.45 | −0.27 | 0.15 | 0.78 |  |
| G1 | Caudate | −1.33 | −0.24 | 0.19 | 0.78 |  |
| G1 | Thalamus | −1.22 | −0.22 | 0.23 | 0.78 |  |
| G2 | Pallidum | −1.08 | −0.20 | 0.28 | 0.78 |  |
| G1 | Hippocampus | 0.90 | 0.17 | 0.37 | 0.78 |  |
| G2 | Putamen | −0.86 | −0.16 | 0.39 | 0.78 |  |
| G2 | Hippocampus | −0.40 | −0.07 | 0.69 | 0.97 |  |
| G1 | Putamen | −0.38 | −0.07 | 0.71 | 0.97 |  |
| G1 | Accumbens | −0.37 | −0.07 | 0.71 | 0.97 |  |
| G1 | Pallidum | 0.30 | 0.06 | 0.76 | 0.97 |  |
| G2 | Amygdala | 0.12 | 0.02 | 0.91 | 0.99 |  |
| G2 | Thalamus | 0.04 | 0.01 | 0.97 | 0.99 |  |
| G2 | Caudate | −0.01 | 0.00 | 0.99 | 0.99 |  |
| **HC vs PT (combined patients)** |  |  |  |  |  |  |
| G1 | Amygdala | −2.83 | −0.42 | 0.01 | 0.07 |  |
| G2 | Accumbens | −1.96 | −0.29 | 0.05 | 0.21 |  |
| G2 | Pallidum | −1.92 | −0.29 | 0.06 | 0.21 |  |
| G2 | Putamen | −1.86 | −0.28 | 0.06 | 0.21 |  |
| G1 | Caudate | −1.79 | −0.27 | 0.08 | 0.21 |  |
| G1 | Accumbens | −1.38 | −0.20 | 0.17 | 0.40 |  |
| G1 | Thalamus | −1.15 | −0.17 | 0.25 | 0.45 |  |
| G1 | Putamen | −1.14 | −0.17 | 0.26 | 0.45 |  |
| G2 | Amygdala | 0.98 | 0.14 | 0.33 | 0.51 |  |
| G1 | Hippocampus | −0.58 | −0.09 | 0.56 | 0.75 |  |
| G1 | Pallidum | 0.46 | 0.07 | 0.65 | 0.75 |  |
| G2 | Hippocampus | −0.42 | −0.06 | 0.67 | 0.75 |  |
| G2 | Thalamus | −0.33 | −0.05 | 0.74 | 0.75 |  |
| G2 | Caudate | 0.32 | 0.05 | 0.75 | 0.75 |  |
| **SZ vs BD** |  |  |  |  |  |  |
| G1 | Hippocampus | 2.67 | 0.61 | 0.01 | 0.13 | ‡ |
| G1 | Amygdala | 1.38 | 0.31 | 0.17 | 0.73 |  |
| G1 | Accumbens | 1.22 | 0.28 | 0.22 | 0.73 |  |
| G2 | Amygdala | -1.09 | -0.25 | 0.28 | 0.73 |  |
| G2 | Putamen | 1.06 | 0.24 | 0.29 | 0.73 |  |
| G2 | Accumbens | -0.96 | -0.22 | 0.34 | 0.73 |  |
| G1 | Putamen | 0.91 | 0.21 | 0.36 | 0.73 |  |
| G2 | Pallidum | 0.77 | 0.18 | 0.45 | 0.79 |  |
| G1 | Thalamus | -0.52 | -0.12 | 0.61 | 0.82 |  |
| G2 | Thalamus | 0.52 | 0.12 | 0.61 | 0.82 |  |
| G2 | Caudate | -0.46 | -0.11 | 0.65 | 0.82 |  |
| G2 | Hippocampus | -0.18 | -0.04 | 0.86 | 0.91 |  |
| G1 | Caudate | 0.12 | 0.03 | 0.90 | 0.91 |  |
| G1 | Pallidum | -0.11 | -0.02 | 0.91 | 0.91 |  |

**Supplementary Table S2. Gradient projection scores statistics across diagnostic contrasts.** Per-region statistics for G1 and G2 gradient projection scores onto six bilateral subcortical structures and hippocampi, comparing healthy controls against schizophrenia, bipolar disorder, and the combined patient group. Negative d indicates lower scores in the comparison group than in HC. Significance markers in the right column: * p_FDR_ < 0.05; ‡ p_unc_ < 0.05. Abbreviations: BD, bipolar disorder; d, Cohen's d effect size; FDR, false discovery rate; G1, first gradient; G2, second gradient; HC, healthy controls; PT, combined patient group; SZ, schizophrenia; t, two-sample t statistic.

##### PLS Out-of-Sample Generalization

For the cortical PLS, both gradient-specific models showed significant out-of-sample generalization. For G1-LV1, the training-set Pearson correlation between brain and behavioral latent scores was r = 0.65 (p < 0.001), with a test-set correlation of r = 0.48 (p = 0.01). For G2-LV1, generalization was comparably strong (train r = 0.74, p < 0.001; test r = 0.41, p = 0.03). The moderate attenuation from training to test partitions is consistent with expected shrinkage under cross-validation and does not indicate overfitting, as evidenced by the significant test-set correlations (**Supplementary Figure S1A**).

Replacing the 400-parcel cortical brain matrix with the seven bilateral gradient projection scores produced markedly weaker out-of-sample generalization. For the subcortical G1-LV1, train r = 0.38 (p = 0.02) and test r = 0.29 (p = 0.08); for the subcortical G2-LV1, train r = 0.33 (p = 0.04) and test r = 0.22 (p = 0.21). In both cases, the test-set correlation failed to reach statistical significance, in contrast to the cortical PLS (**Supplementary Figure S1B**). This pattern indicates that the brain–behaviour covariance structure identified in the primary analyses is carried predominantly by cortical gradient organization and is not recoverable from connectivity-weighted gradient projection scores alone, a finding consistent with the comparatively circumscribed univariate subcortical effects (restricted to the amygdala along G1) reported in the main text.

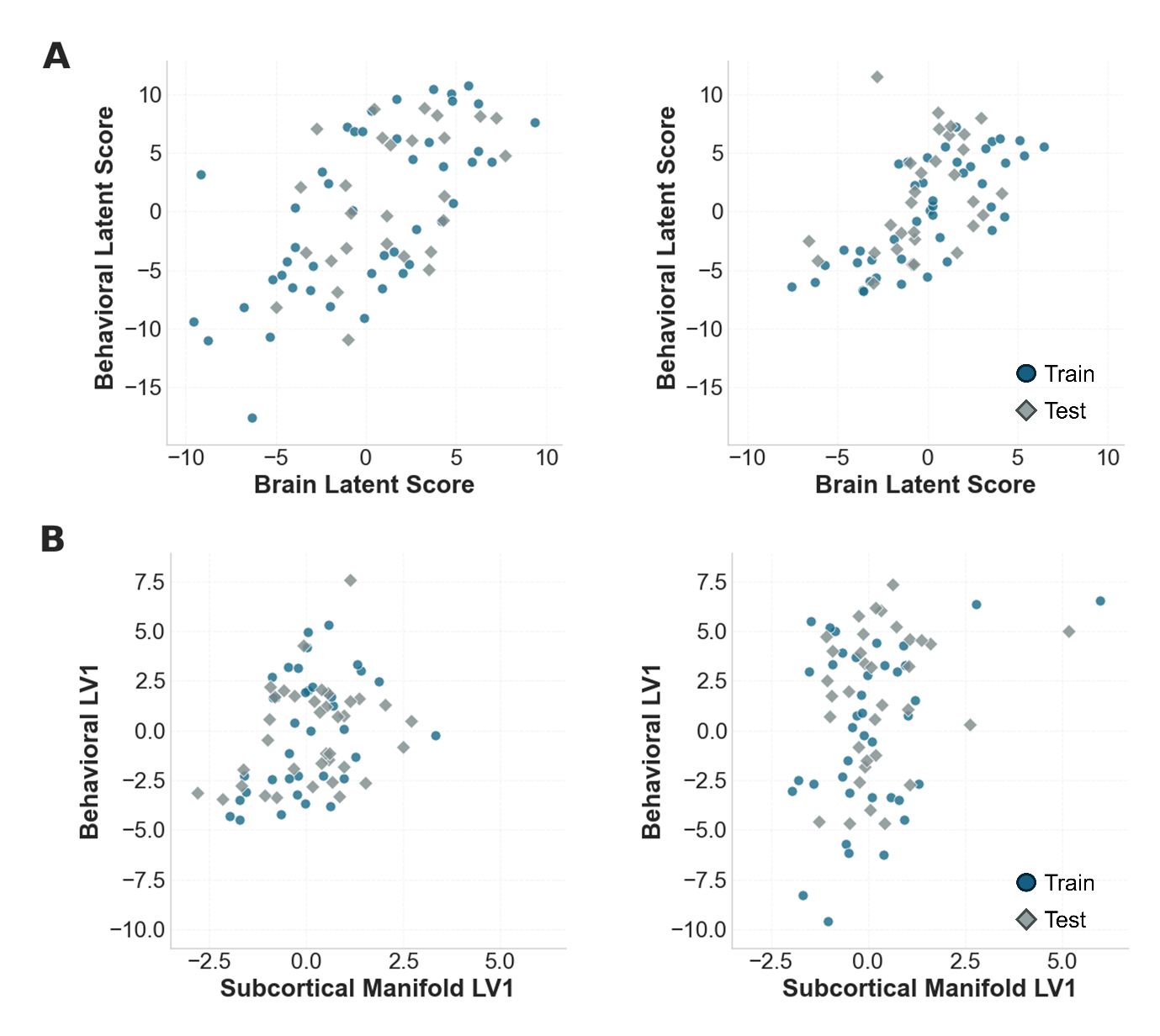

**Supplementary Figure S1. Out-of-sample generalization of cortical and subcortical PLS models.** **A.** Cortical PLS. Scatter plots of brain latent scores (X · u) against behavioral latent scores (Y · v) for G1-LV1 (left; train r = 0.65, p < .001; test r = 0.48, p = 0.011) and G2-LV1 (right; train r = 0.74, p < .001; test r = 0.41, p = 0.026). Each point represents one patient; teal circles indicate the training partition and grey diamonds the held-out test partition. **B.** Subcortical PLS. Equivalent scatter plots for G1-LV1 (left; train r = 0.38, p = 0.02; test r = 0.29, p = 0.08) and G2-LV1 (right; train r = 0.33, p = 0.04; test r = 0.22, p = 0.21).

##### Spatial Correspondence Analysis: Network-Specific Contributions

At the network level, the whole-brain G1 null result concealed opposing network-specific effects that cancelled across the cortex: within visual cortex, PLS salience positively followed the G1 gradient topology (r = +0.40, p_FDR_ = 0.011), whereas within the dorsal attention network it was inverted (r = −0.46, p_FDR_= 0.011), indicating that the G1 brain-behaviour signal reflects a selective and directionally dissociated reorganization rather than a uniform axis shift. For G2, the frontoparietal network showed the strongest inversion (r = −0.36, p_FDR_= 0.007); salience/ventral attention and somatomotor networks showed consistent negative trends that did not survive FDR correction.

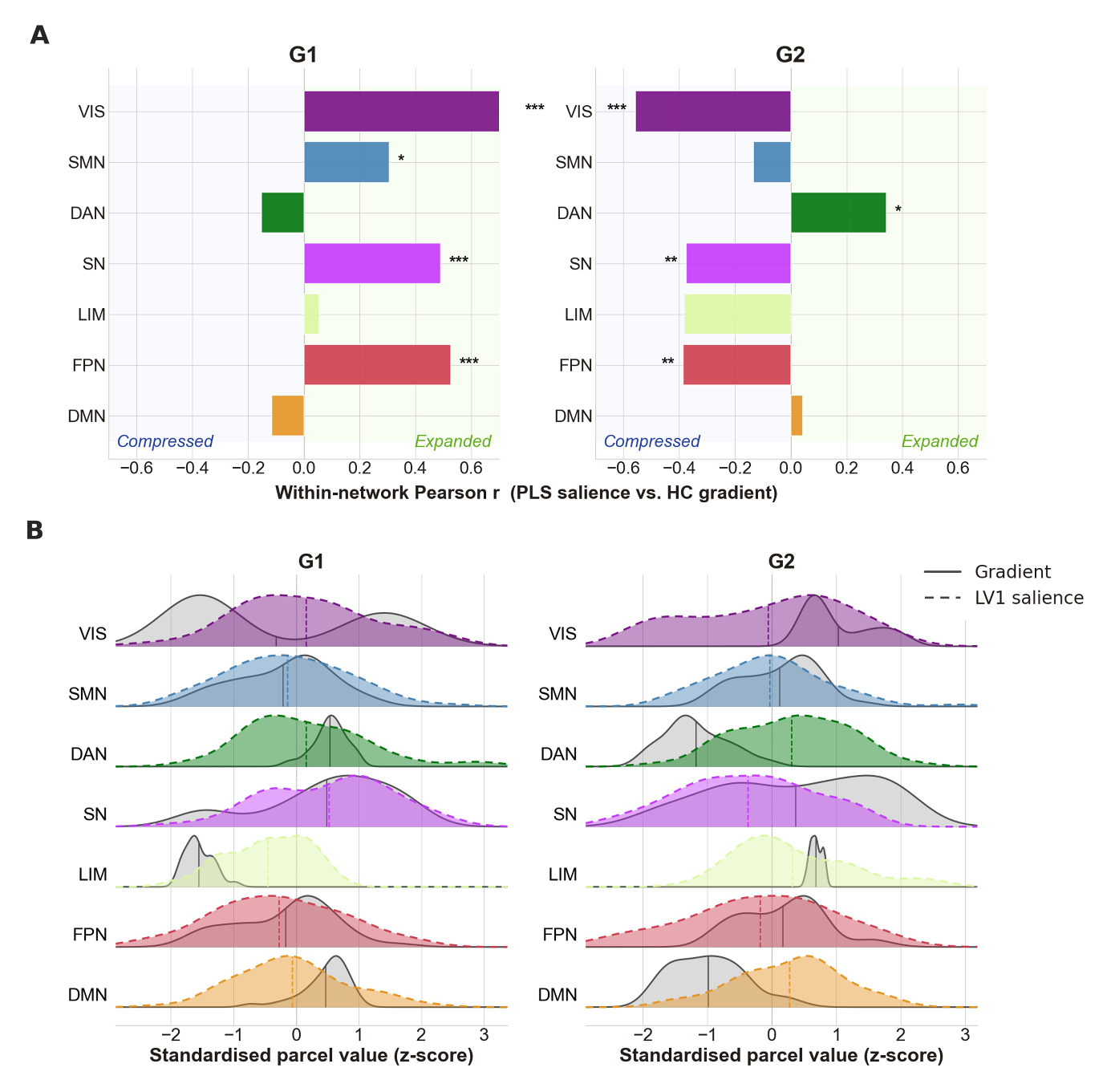

**Supplementary Figure S2. Network-specific spatial correspondence between PLS brain salience and the normative cortical gradient. A.** Within-network Pearson correlations between the PLS-LV1 brain salience map and the healthy-control (HC) group-mean gradient map, computed separately for each of the seven Schaefer functional networks, for G1 (left) and G2 (right). Asterisks indicate FDR-corrected significance (* p_FDR_ < 0.05; ** p_FDR_< 0.01; *** p_FDR_< 0.001). **B.** Ridge plot showing, for each Schaefer network (rows), the parcel-level density of the normative gradient values (solid grey ridges) and of the PLS LV1 brain salience weights (dashed, network-colored ridges). Both distributions are z-scored within each panel, the vertical line marks the panel-wide mean (z = 0). Abbreviations: VIS, visual; SMN, somatomotor; DAN, dorsal attention; SN, salience; LIM, limbic; FPN, frontoparietal; DMN, default mode network.

##### Gradient Similarity Index

Within the combined patient sample, SI-PLS scores showed robust and dissociated associations with symptoms and cognitive variables. For G1-SI-PLS, higher alignment with the G1 BD-associated salience pattern was associated with lower psychopathology burden across positive symptom severity (r = −0.50, p_FDR_ < 0.001), diminished expression (r = −0.44, p_FDR_< 0.001), disorganization (r = −0.32, p_FDR_= 0.014), and amotivation (r = −0.33, p_FDR_= 0.014). For G2-SI-PLS, higher alignment with the G2 SZ-associated salience pattern was associated with greater positive symptom severity (r = 0.41, p_FDR_= 0.001), diminished expression (r = 0.41, p_FDR_= 0.001), amotivation (r = 0.33, p_FDR_= 0.01), and worse cognitive performance in attention/working memory (r = 0.29, p_FDR_= 0.02), verbal learning (r = 0.35, pFDR = 0.004), and vocabulary (r = 0.36, p_FDR_= 0.004). Mood symptoms (depression, mania) and reasoning were not significantly associated with either index (all p_FDR_> 0.07). The full pattern of correlations is shown in **Supplementary Figure S3** and visualizes the diagnostic dissociation between the two gradient-derived indices: G1-SI-PLS loaded negatively across psychopathology domains (BD-leaning profile), whereas G2-SI-PLS loaded positively across both symptom and cognitive domains (SZ-leaning profile).

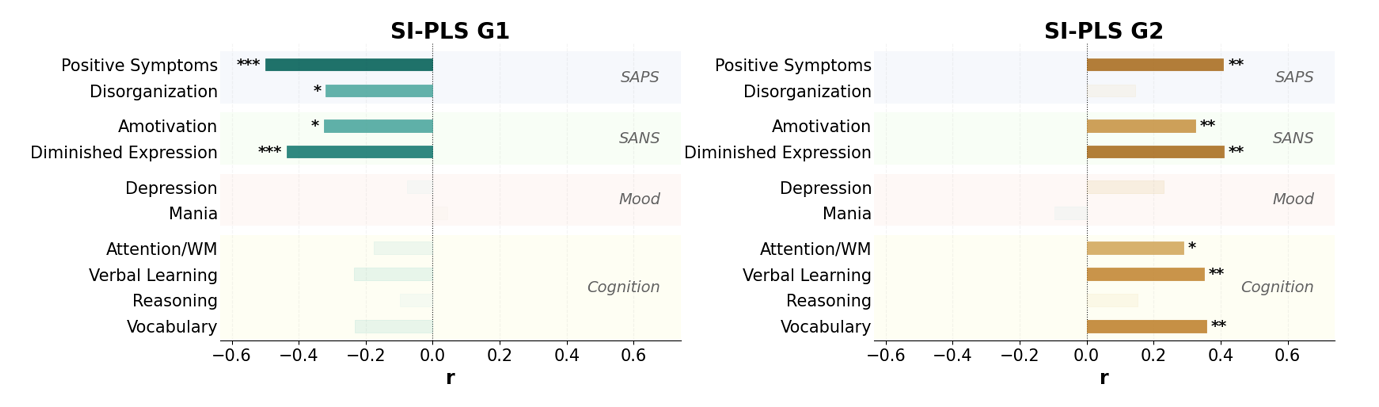

**Supplementary Figure S3. Clinical correlations of the PLS-derived similarity index (SI-PLS).** Pearson correlations between SI-PLS scores and behavioral variables within the combined patient sample (N = 77), separately for G1-SI-PLS (left) and G2-SI-PLS (right). Bars represent Pearson r; saturated colors denote FDR-significant associations, faded bars indicate non-significant correlations. Asterisks indicate FDR-corrected significance (* p_FDR_ < 0.05; ** p_FDR_ < 0.01; *** p_FDR_ < 0.001). Abbreviations: WM, working memory; SAPS, Scale for the Assessment of Positive Symptoms; SANS, Scale for the Assessment of Negative Symptoms.
